# Risk of Emergent Dolutegravir Resistance Mutations In People Living With HIV: A Rapid Scoping Review

**DOI:** 10.1101/2024.01.05.24300911

**Authors:** Carolyn Chu, Kaiming Tao, Vinie Kouamou, Ava Avalos, Jake Scott, Philip M. Grant, Soo Yon Rhee, Suzanne M. McCluskey, Michael R. Jordan, Rebecca L. Morgan, Robert W. Shafer

## Abstract

**Background:** Dolutegravir (DTG) is a cornerstone of global antiretroviral (ARV) therapy (ART) due to its high efficacy and favorable tolerability. However, limited data exist regarding the risk of virological failure (VF) with emergent integrase strand transfer inhibitor (INSTI) drug-resistance mutations (DRMs) in individuals receiving DTG-containing ART.

**Methods:** We performed a PubMed search using the term “Dolutegravir” last updated December 18, 2023, to estimate the prevalence of VF with emergent INSTI DRMs in clinical trials and cohorts of people living with HIV (PLWH) without previous VF on an INSTI who received DTG-containing ART.

**Results:** Of 2131 records identified by search, 43 clinical trials, 39 cohorts, and six cross-sectional studies provided data across six clinical scenarios based upon ART history, virological status, and ARVs co-administered with DTG: (1) ART-naïve PLWH receiving DTG plus two nucleoside reverse transcriptase inhibitors (NRTIs); (2) ART-naïve PLWH receiving DTG plus lamivudine; (3) ART-experienced PLWH with VF on their previous regimen receiving DTG plus two NRTIs; (4) ART-experienced PLWH with virological suppression receiving DTG plus two NRTIs; (5) ART-experienced PLWH with virological suppression receiving DTG and a second ARV; and (6) ART-experienced PLWH with virological suppression receiving DTG monotherapy. The proportion of PLWH in clinical trials with emergent INSTI DRMs was 1.6% for scenario 3 and 2.9% for scenario 6. In the remaining four trial scenarios, prevalence of VF with emergent INSTI DRMs was ≤0.1%. Data from cohort studies minimally influenced prevalence estimates from clinical trials, while cross-sectional studies yielded prevalence data lacking denominator details.

**Conclusions:** In clinical trials, the prevalence of VF with emergent INSTI DRMs in PLWH receiving DTG-containing regimens has been low. Novel approaches are required to assess the risk of VF with emergent INSTI DRMs in PLWH receiving DTG in real-world settings.

**What is already known on this topic:** Dolutegravir is known for its high resistance barrier, yet there remains a concern for virological failure and subsequent drug resistance in people living with HIV who begin first or second-line antiretroviral therapy with a dolutegravir-containing regimen.

**What this study adds:** The prevalence of virological failure with the development of HIV mutations associated with reduced susceptibility to dolutegravir depends on a person’s virological response to previous antiretroviral therapy, the presence of HIV replication at dolutegravir initiation, and the antiretroviral drugs co-administered with dolutegravir.

In clinical trial settings, the prevalence of virological failure with emergent dolutegravir resistance was rare among people initiating therapy with a dolutegravir-containing regimen and was 1.6% over a period of one to two years among those who had previously experienced virological failure on an earlier treatment regimen.

In the subset of persons with virological failure on a first-line dolutegravir-containing regimen, the prevalence of emergent dolutegravir resistance was 0.7%, whereas in the subset of persons with virological failure on a second-line dolutegravir-containing regimen, the prevalence of emergent dolutegravir resistance was 20.4%.

**How this study might affect research, practice, or policy:** In people living with HIV with virological failure on a first-line dolutegravir-containing regimen, enhancing medication adherence may prove more beneficial than transitioning to an alternative treatment regimen.

In cases of virological failure on a second-line dolutegravir-containing regimen, the potential for dolutegravir resistance suggests a need to investigate the role of genotypic resistance testing to inform treatment changes.

Population-level surveillance for acquired dolutegravir resistance should take into account the antiretroviral treatment history and level of HIV replication prior to the initiation of dolutegravir-containing therapy.

## INTRODUCTION

Among integrase strand transfer inhibitor (INSTI)-naïve people living with HIV (PLWH), antiretroviral (ARV) therapy (ART) with the second-generation INSTIs dolutegravir (DTG) or bictegravir (BIC) has been associated with low rates of virological failure (VF) and emergent INSTI-associated drug-resistance mutations (DRMs). Of these two INSTIs, DTG is instrumental in the World Health Organization’s (WHO’s) efforts to improve global viral suppression rates because DTG-containing regimens are recommended in multiple clinical scenarios, including for first-line ART, programmatic transition for individuals being treated with a first-line nonnucleoside reverse transcriptase inhibitor (NNRTI)-based regimen, and second-line ART following VF on a first-line NNRTI-based regimen [1]. By 2022, an estimated 22.2 million PLWH in low- and middle-income countries (LMICs) were receiving a DTG-containing regimen [2].

We recently published a systematic review that identified 36 publications reporting 99 INSTI-naïve PLWH with emergent VF and INSTI DRMs on a DTG-containing regimen [3]. However, this review did not estimate the risk of emergent INSTI DRMs in INSTI-naïve PLWH because most individuals with INSTI DRMs were reported in studies for which the total number of PLWH receiving DTG was unknown. Additionally, this review’s search identified only studies reporting INSTI DRMs and not those in which DTG was received but INSTI DRMs were not reported.

Therefore, we perform here a scoping review to estimate the risk of VF with emergent INSTI DRMs in PLWH without a history of VF on a previous INSTI-based regimen according to (1) whether they were ART-naïve or experienced, (2) were virologically suppressed when DTG-containing ART was initiated, and (3) the ARVs co-administered with DTG. Our review included clinical trials, observational cohort studies, and cross-sectional studies in which PLWH underwent genotypic resistance testing (GRT). We confined our analysis to PLWH without a history of VF on a first-generation INSTI because such VF is often associated with DTG cross-resistance or a marked reduction in the genetic barrier to DTG resistance [4]. Additionally, first-generation INSTIs have rarely been used in LMICs, where the concern about emergent DTG resistance is the greatest.

## METHODS

This rapid scoping review evaluated the risk of VF with emergent INSTI DRMs in PLWH without VF on a first-generation INSTI. The review adhered to the Preferred Reporting in Systematic Review and Meta-Analysis (PRISMA) extension for Scoping Reviews [5]. One author (RWS) reviewed the results of a PubMed query using the search term “Dolutegravir” from database inception through December 18, 2023, to identify clinical trials, cohort studies, and cross-sectional studies containing data on the prevalence of emergent INSTI DRMs in populations without previous VF on an INSTI-containing regimen who received a DTG-containing regimen. We did not involve patients or the public in the design, or conduct, or reporting, or dissemination plans of our research.

Publications were excluded for any of the following reasons: (i) absence of GRT data; (ii) inclusion of individuals with previous VF on a first-generation INSTI whose data could not be distinguished from those who were previously INSTI-naïve; (iii) lack of essential information such as ART history, viral replication status at the initiation of DTG, or the ARVs co-administered with DTG; (iv) studies with fewer than 30 individuals; (v) studies not reporting individual-level VF data; (vi) studies with data previously reported in an already included study; or (vii) studies not involving individuals treated with DTG.

Clinical trials, cohort studies, and cross-sectional studies that included PLWH who had previously received a first-generation INSTI were eligible only if the study subjects had been virologically suppressed at the time DTG-containing ART was begun and had not previously experienced VF on an INSTI-containing regimen. Studies that reported no instances of VF were included despite the fact that GRT was not performed.

The following data were extracted from published studies meeting eligibility criteria: (1) geographic region; (2) study population characteristics including age range, pregnancy, and whether rifampin-containing antituberculosis (anti-TB) treatment was co-administered; (3) ART history; (4) whether virological suppression (VS) was present at the time DTG-containing ART was begun; (5) the ARVs co-administered with DTG; (6) the number of individuals receiving a DTG-containing regimen; (7) the proportion of individuals with protocol-defined VF at specific time points such as weeks 24, 48, and 96; (8) the definition of author-defined VF; (9) the proportion of individuals undergoing GRT; and (10) the proportion of individuals with emergent INSTI DRMs. Data extraction was performed primarily by CC and RWS with the assistance of VK, AA, JS, and PMG.

Results are presented as the proportions of PLWH experiencing VF, undergoing GRT, and developing INSTI DRMs. Clinical trials and observational studies meeting search criteria were grouped into six clinical scenarios based on the population’s ART experience, the presence of VS prior to initiating DTG-containing ART, and the ARVs co-administered with DTG.

Complete HIV-1 sequences were rarely reported by authors or submitted to GenBank. Therefore, we relied on the reports of integrase mutations in each published study. INSTI-associated DRMs were defined as the following nonpolymorphic mutations: H51Y, T66A/I/K, E92G/Q, G118R, F121Y, E138A/K/T, G140A/C/S, Y143C/H/R/S, S147G, Q148H/R/K, S153Y/F, N155H, S230R, and R263K [3].

## RESULTS

As of December 18, 2023, 2,131 publications were retrieved from PubMed. Following title and abstract review, 345 publications were selected for full-text review. Of 105 publications describing clinical trials, 32 were excluded because they (i) did not contain GRT data; (ii) included individuals with previous VF on a first-generation INSTI; (iii) contained pharmacokinetic data only; (iv) contained fewer than 30 individuals; or (iv) contained the same information reported in another study (Figure 1).

**Figure 1.**
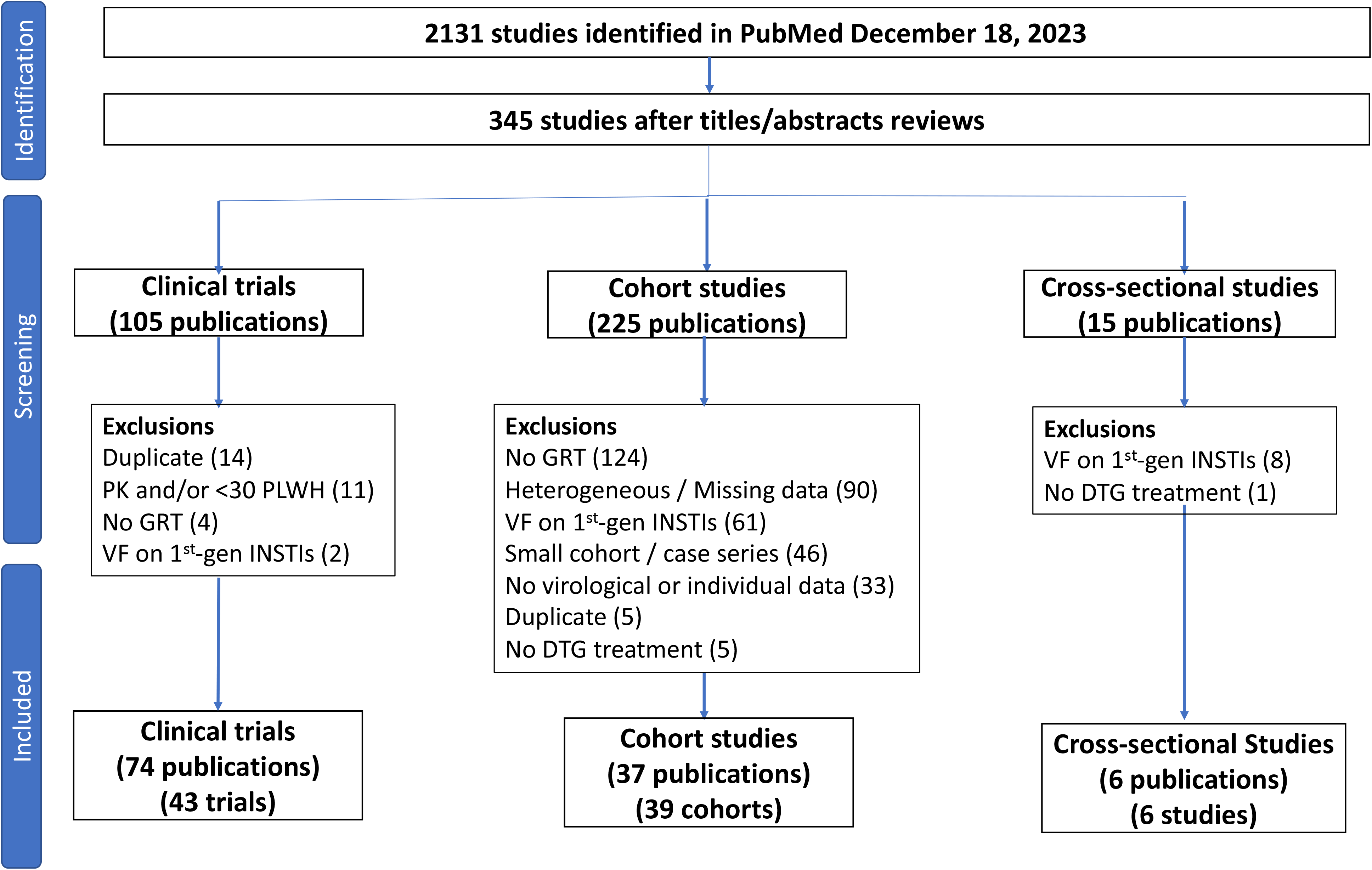
Flow chart summarizing the review process. Of 2131 publications identified in the PubMed search, 345 were read in their entirety following an initial review of titles and abstracts. Exclusions for clinical trials and cross-sectional studies were usually based on a single exclusion criterion. Exclusions for cohort studies were usually based on more than one exclusion criterion. Two publications contained descriptions of two cohorts. Abbreviations: PK – pharmacokinetic study; PLWH – people living with HIV; GRT – genotypic resistance testing; VF – virological failure; 1^st^-gen INSTI – previous VF on a 1^st^-generation integrase strand transfer inhibitor (raltegravir or elvitegravir). Heterogeneous / Missing data – indicates that the study described different subsets of individuals with different ART histories, levels of virus suppression, and DTG-containing regimens but that the virological and/or GRT outcomes were not provided for the different subsets.

Of 225 publications describing clinical cohorts, 186 were excluded because they (i) did not contain GRT data; (ii) were missing critical data such as ART history and virological status (i.e., whether patients were virologically suppressed) prior to DTG initiation; (iii) included individuals with previous VF on a first-generation INSTI; (iv) contained fewer than 30 individuals; (v) did not contain individual-level VF data; (vi) contained the same information reported in another study; and/or (vii) did not include individuals who received DTG (Figure 1).

Of 15 publications describing cross-sectional studies of GRT in PLWH with VF after receiving one or more INSTIs, 9 were excluded because they (i) did not distinguish individuals who experienced VF on DTG alone versus those who received DTG following VF on a first-generation INSTI or (ii) did not include persons receiving a DTG-containing regimen (Figure 1).

Following full-text review, 74 publications describing 43 clinical trials, 37 publications describing 39 cohort studies, and six publications describing cross-sectional studies met study eligibility criteria. When grouped according to population ART history, level of virus replication, and ARVs co-administered with DTG, the studies were classified into six clinical scenarios: (1) ART-naïve PLWH receiving initial therapy with DTG plus two nucleoside RT inhibitors (NRTIs); (2) ART-naïve PLWH receiving initial therapy with DTG plus lamivudine (3TC); (3) ART-experienced PLWH with previous VF on an NNRTI-containing regimen receiving DTG plus two NRTIs or an optimized background regimen; (4) ART-experienced PLWH with VS switching to a regimen of DTG plus two NRTIs; (5) ART-experienced PLWH with VS switching to a two-drug DTG-containing regimen; and (6) ART-experienced PLWH with VS switching to DTG monotherapy. The numbers of clinical trials, cohort studies, and cross-sectional studies describing populations belonging to each of these scenarios is shown in Table 1.

**Table 1.** The Numbers of Clinical Trials, Cohort Studies, and Cross-Sectional Studies Belonging to Each of the Six Clinical Scenarios.

| **Clinical Scenario** | **ART History** | **Viral Load Prior to DTG** | **DTG-Containing ART** | **Clinical Trials (number trials)** | **Clinical Trials (number PLWH)** | **Cohort Studies (number studies)** | **Cohort Studies (number PLWH)** | **Cross-Sectional Studies (number studies)** | **Cross-Sectional Studies (number PLWH)** |
| --- | --- | --- | --- | --- | --- | --- | --- | --- | --- |
| 1 | Naïve | Viremic | DTG + 2 NRTIs | 16 | 4585 | 7 | 2698 | 1 | 113 |
| 2 | Naïve | Viremic | DTG + 3TC | 4 | 1075 | 3 | 581 | 0 | 0 |
| 3 | Experienced | Viremic | DTG + 2 NRTIs | 6 | 1428 | 5 | 3630 | 4 | 218 |
| 4 | Experienced | Suppressed | DTG + 2 NRTIs | 3 | 877 | 3 | 2204 | 0 | 0 |
| 5 | Experienced | Suppressed | DTG + 2<sup>nd</sup> ARV | 10 | 1894 | 18 | 5930 | 0 | 0 |
| 6 | Experienced | Suppressed | DTG monotherapy | 4 | 276 | 3 | 123 | 0 | 0 |

Abbreviations: ARV – antiretroviral; ART – antiretroviral therapy; DTG – dolutegravir; PLWH – people living with HIV

### ART-Naïve PLWH

#### DTG plus two NRTIs

Table 2 summarizes the population characteristics, prevalence of VF, and GRT results in 16 clinical trials of 4636 ART-naïve PLWH treated with DTG plus two NRTIs [6–34]. The trials included seven multicenter registration trials in which participants were evaluated at week 96 (and week 144 in one trial), two trials from Sub-Saharan Africa in which participants were evaluated at week 96, two trials in which participants were evaluated at week 48, and five trials of special populations. The latter trials included PLWH who were also receiving anti-TB therapy, pregnant women, individuals with acute HIV-1 infection, and individuals co-infected with hepatitis B virus. VF was defined as a confirmed plasma HIV-1 RNA level ≥50 copies/ml in eight trials, ≥200 copies/ml in three trials, ≥400 copies/ml in three trials, and ≥1000 copies/ml in two trials.

**Table 2.**
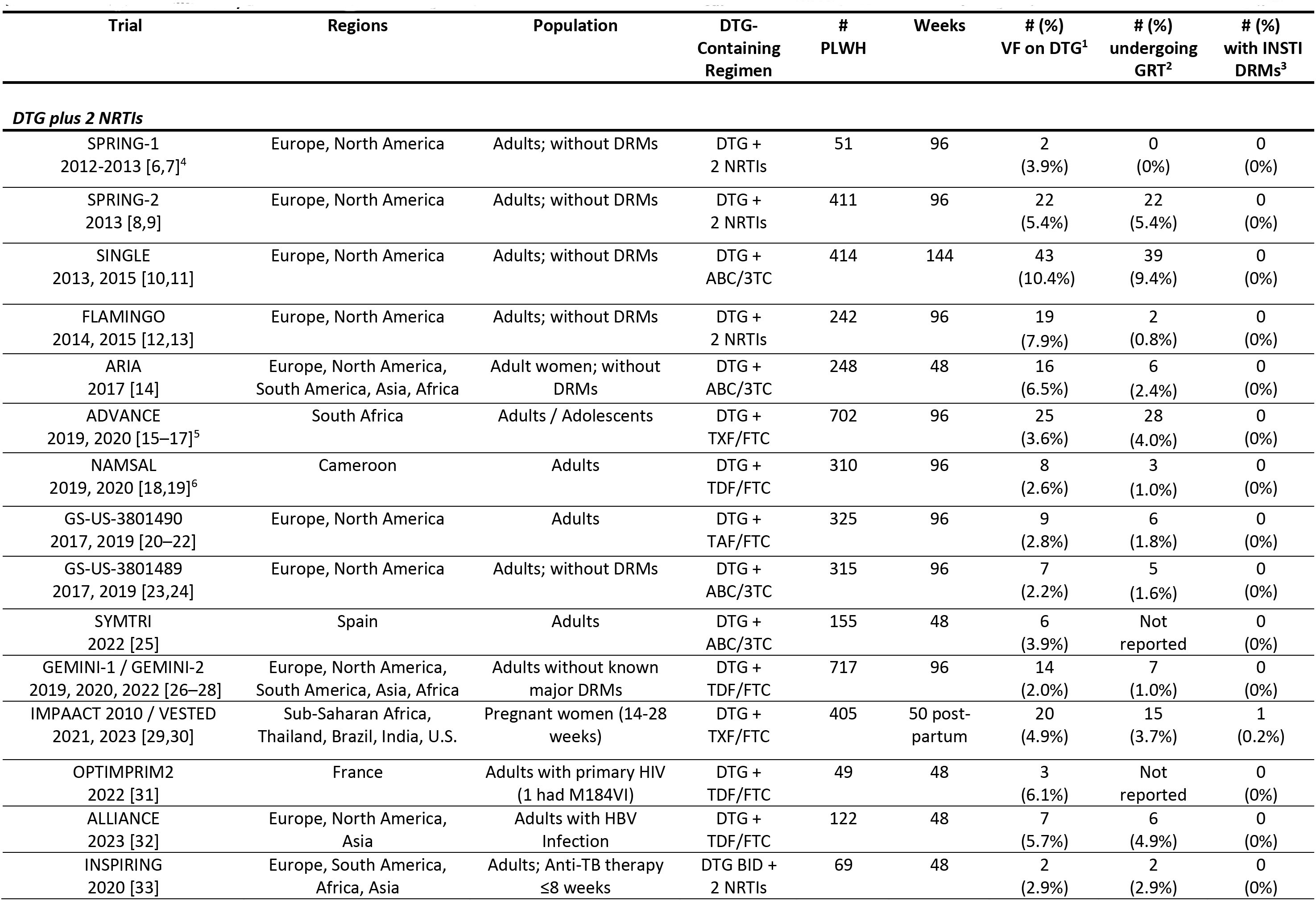
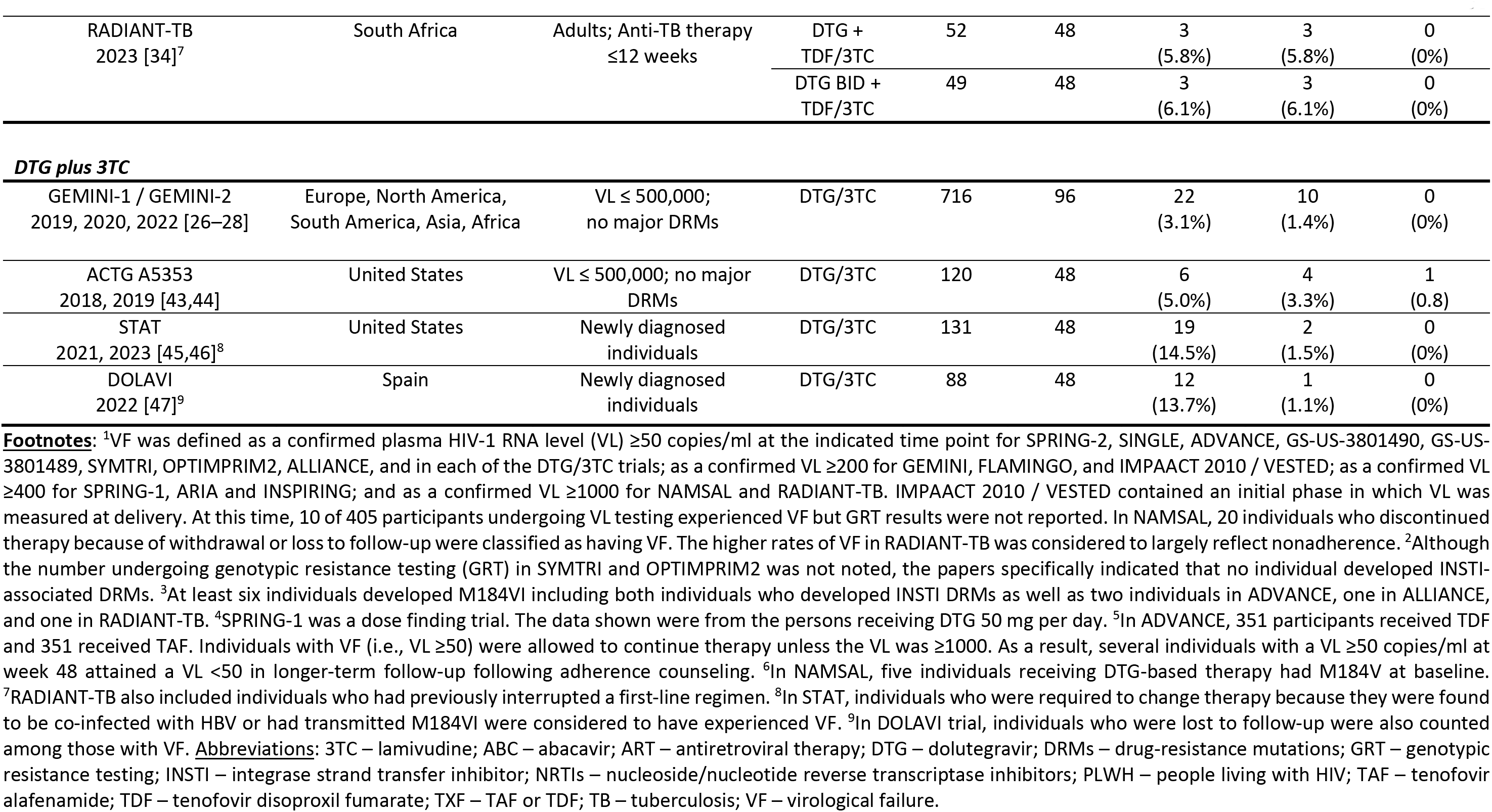
Virological Failure (VF) and Prevalence of Emergent INSTI-Associated DRMs in Clinical Trials of ART-Naïve PLWH Receiving a First-Line DTG-Containing Regimen.

| Trial | Regions | Population | DTG-Containing Regimen | # PLWH | Weeks | # (%) VF on DTG<sup>1</sup> | # (%) undergoing GRT<sup>2</sup> | # (%) with INSTI DRMs<sup>3</sup> |
| --- | --- | --- | --- | --- | --- | --- | --- | --- |
| ***DTG plus 2 NRTIs*** |  |  |  |  |  |  |  |  |
| SPRING-1<br>2012-2013 [6,7]<sup>4</sup> | Europe, North America | Adults; without DRMs | DTG + 2 NRTIs | 51 | 96 | 2 (3.9%) | 0 (0%) | 0 (0%) |
| SPRING-2<br>2013 [8,9] | Europe, North America | Adults; without DRMs | DTG + 2 NRTIs | 411 | 96 | 22 (5.4%) | 22 (5.4%) | 0 (0%) |
| SINGLE<br>2013, 2015 [10,11] | Europe, North America | Adults; without DRMs | DTG + ABC/3TC | 414 | 144 | 43 (10.4%) | 39 (9.4%) | 0 (0%) |
| FLAMINGO<br>2014, 2015 [12,13] | Europe, North America | Adults; without DRMs | DTG + 2 NRTIs | 242 | 96 | 19 (7.9%) | 2 (0.8%) | 0 (0%) |
| ARIA<br>2017 [14] | Europe, North America, South America, Asia, Africa | Adult women; without DRMs | DTG + ABC/3TC | 248 | 48 | 16 (6.5%) | 6 (2.4%) | 0 (0%) |
| ADVANCE<br>2019, 2020 [15–17]<sup>5</sup> | South Africa | Adults / Adolescents | DTG + TXF/FTC | 702 | 96 | 25 (3.6%) | 28 (4.0%) | 0 (0%) |
| NAMSAL<br>2019, 2020 [18,19]<sup>6</sup> | Cameroon | Adults | DTG + TDF/FTC | 310 | 96 | 8 (2.6%) | 3 (1.0%) | 0 (0%) |
| GS-US-3801490<br>2017, 2019 [20–22] | Europe, North America | Adults | DTG + TAF/FTC | 325 | 96 | 9 (2.8%) | 6 (1.8%) | 0 (0%) |
| GS-US-3801489<br>2017, 2019 [23,24] | Europe, North America | Adults; without DRMs | DTG + ABC/3TC | 315 | 96 | 7 (2.2%) | 5 (1.6%) | 0 (0%) |
| SYMTRI<br>2022 [25] | Spain | Adults | DTG + ABC/3TC | 155 | 48 | 6 (3.9%) | Not reported | 0 (0%) |
| GEMINI-1 / GEMINI-2<br>2019, 2020, 2022 [26–28] | Europe, North America, South America, Asia, Africa | Adults without known major DRMs | DTG + TDF/FTC | 717 | 96 | 14 (2.0%) | 7 (1.0%) | 0 (0%) |
| IMPAACT 2010 / VESTED<br>2021, 2023 [29,30] | Sub-Saharan Africa, Thailand, Brazil, India, U.S. | Pregnant women (14-28 weeks) | DTG + TXF/FTC | 405 | 50 post-partum | 20 (4.9%) | 15 (3.7%) | 1 (0.2%) |
| OPTIMPRIM2<br>2022 [31] | France | Adults with primary HIV (1 had M184VI) | DTG + TDF/FTC | 49 | 48 | 3 (6.1%) | Not reported | 0 (0%) |
| ALLIANCE<br>2023 [32] | Europe, North America, Asia | Adults with HBV Infection | DTG + TDF/FTC | 122 | 48 | 7 (5.7%) | 6 (4.9%) | 0 (0%) |
| INSPIRING<br>2020 [33] | Europe, South America, Africa, Asia | Adults; Anti-TB therapy ≤8 weeks | DTG BID + 2 NRTIs | 69 | 48 | 2 (2.9%) | 2 (2.9%) | 0 (0%) |
| RADIANT-TB<br>2023 [34]<sup>7</sup> | South Africa | Adults; Anti-TB therapy<br>≤12 weeks | DTG +<br>TDF/3TC | 52 | 48 | 3<br>(5.8%) | 3<br>(5.8%) | 0<br>(0%) |
|  |  |  | DTG BID +<br>TDF/3TC | 49 | 48 | 3<br>(6.1%) | 3<br>(6.1%) | 0<br>(0%) |
| **DTG plus 3TC** |  |  |  |  |  |  |  |  |
| GEMINI-1 / GEMINI-2<br>2019, 2020, 2022 [26–28] | Europe, North America,<br>South America, Asia, Africa | VL ≤ 500,000;<br>no major DRMs | DTG/3TC | 716 | 96 | 22<br>(3.1%) | 10<br>(1.4%) | 0<br>(0%) |
| ACTG A5353<br>2018, 2019 [43,44] | United States | VL ≤ 500,000; no major<br>DRMs | DTG/3TC | 120 | 48 | 6<br>(5.0%) | 4<br>(3.3%) | 1<br>(0.8%) |
| STAT<br>2021, 2023 [45,46]<sup>8</sup> | United States | Newly diagnosed<br>individuals | DTG/3TC | 131 | 48 | 19<br>(14.5%) | 2<br>(1.5%) | 0<br>(0%) |
| DOLAVI<br>2022 [47]<sup>9</sup> | Spain | Newly diagnosed<br>individuals | DTG/3TC | 88 | 48 | 12<br>(13.7%) | 1<br>(1.1%) | 0<br>(0%) |

The median proportion of PLWH experiencing VF in the 16 trials was 4.9% (IQR: 2.8%-6.1% range: 2.0%-10.4%). At the time of VF, GRT was performed on 147 individuals (3.2% of all participants) receiving DTG and only one (0.02% of all participants) developed an INSTI DRM. This individual was one of 405 women treated during the second trimester of pregnancy and evaluated 50 weeks post-partum [30]. The emergent INSTI DRMs included S147G, N155H, and S230R. This individual and at least four other individuals acquired the 3TC/emtricitabine (FTC)- associated reverse transcriptase (RT) mutation, M184VI.

Seven cohort studies described 2698 ART-naïve PLWH treated with DTG plus two NRTIs including two studies from the U.S. [35,36], two from Italy [37,38], and one each from Spain [39], South Korea [40], and Tanzania [41] (Supplementary Table 1). One of the two studies from the U.S. included just pregnant women [35]. The median duration of follow-up ranged from nine to 26 months, and the proportions with VF ranged from 0% to 23%. None of these studies reported new instances of emergent INSTI DRMs.

One cross-sectional study from Brazil reported that among 113 previously ART-naïve PLWH with confirmed VF while receiving DTG plus tenofovir disoproxil fumarate (TDF)/3TC, seven (6.2%) individuals had major INSTI DRMs [42] (Supplementary Table 1). However, the size of the population receiving DTG plus TDF/3TC and the total number with VF was not reported in this study.

#### DTG plus 3TC

Two phase 3, one phase 2, and one open-label non-randomized clinical trial examined the prevalence of VF with emergent INSTI DRMs in ART-naïve individuals receiving a first-line DTG plus 3TC regimen (Table 2) [26–28,43–48]. In each trial, VF was defined as either a single or confirmed VL ≥50 copies/ml at the specified time point. Of the 1055 PLWH in these trials, 59 (5.6%) experienced VF and 17 (1.6%) underwent GRT by week 48 or 96. One individual (0.1% of the total) developed an INSTI DRM, R263K, in combination with the 3TC-associated DRM M184VI.

Three cohort studies included 581 PLWH who received DTG plus 3TC and were followed for a median of 48 weeks [39,49,50] (Supplementary Table 2). Two studies were from Spain, and one was from China. Four individuals (0.7%) were considered to have VF. GRT was performed on samples from two individuals (0.3%) across the studies, and neither individual had an INSTI DRM.

### ART-Experienced PLWH with a History of VF on an NNRTI-Containing Regimen

Table 3 summarizes data from five clinical trials of DTG plus two NRTIs and one trial of DTG plus an optimized background regimen for treating PLWH with a history of VF on a previous NNRTI-containing regimen [51–64]. Four trials were conducted in adults and/or adolescents [51–59] and two in infants, children, and/or adolescents [60–64]. VF was defined as a plasma HIV-1 RNA level ≥50 copies/ml at week 24 in the ARTIST trial and ≥400 copies/ml at weeks 48 or 96 in the remaining trials.

**Table 3.** Virological Failure (VF) and Prevalence of Emergent INSTI DRMs in Clinical Trials of ART-Experienced PLWH with Active Virus Replication Receiving DTG Plus Two 2 NRTIs.

| Trial | Regions | Population<sup>1</sup> | DTG-Containing Regimen<sup>2</sup> | # PLWH | Weeks | # (%) VF<sup>3</sup> | # (%) Undergoing GRT | # (%) with INSTI DRMs<sup>4</sup> |
| --- | --- | --- | --- | --- | --- | --- | --- | --- |
| SAILING<br>2013, 2015 [51,52] | Europe, North America, South America, Asia, Oceania, Africa | Adults; VL≥400; INSTI-naïve, ≥2-class resistance | DTG + OBR | 354 | 48 | 21 (5.9%) | 9 (2.5%) | 2 (0.6%) |
| DAWNING<br>2019, 2022 [53,54] | Europe, South America, Asia, Africa | Adults; VL≥400 on a first-line NNRTI-containing regimen | DTG + 2 NRTIs (≥1 fully active) | 312 | 48 | 11 (3.5%) | 11 (3.5%) | 3 (1.0%) |
| NADIA<br>2021, 2022 [55,56]<sup>5</sup> | Uganda, Kenya, Zimbabwe | Adults/Adolescents; VL≥1000 on a first-line NNRTI-containing regimen | DTG + TDF/3TC or AZT/3TC | 235 | 96 | 24 (10.2%) | 21 (8.9%) | 9 (3.8%) |
| ARTIST<br>2021, 2023 [57–59] | South Africa | Adults; VL≥400 while on a first-line NNRTI-containing regimen | DTG + TDF/3TC | 135 | 24 | 21 (15.6%) | 4 (3.0%) | 0 (0%) |
|  |  |  | DTG BID + TDF/3TC | 64 | 24 | 9 (14.1%) | 3 (4.7%) | 0 (0%) |
| IMPACT P1093<br>2015, 2020, 2022 [60–62] | North America, South America, Asia, Africa | Infants/children/ adolescents with VL≥1000; most ART-experienced | DTG + 2 NRTIs | 142 | 48 | 36 (25.3%) | 36 (25.3%) | 5 (3.5%) |
| ODYSSEY<br>2021, 2022 [63,64] | Europe, Asia, Africa | Children and adolescents with V ≥500 on a 1<sup>st</sup>- or 2<sup>nd</sup>-line ART regimen | DTG + 2 NRTIs | 196 | 96 | 31 (15.8%) | 29 (14.7%) | 4 (2.0%) |

In the SAILING trial, baseline GRT was used to select the ARVs co-administered with DTG to create an optimized background regimen. In the DAWNING trial, baseline GRT was used to exclude individuals in whom no NRTI was predicted to retain antiviral activity. By contrast, in the remaining trials, baseline GRT results were neither available in real time nor used to guide therapy. Retrospective analysis of baseline samples in the NADIA and ARTIST trials indicated high levels of baseline NRTI-associated DRMs. In the NADIA trial, approximately 85% of baseline samples had the 3TC-resistance mutations M184VI while about 50% had a TDF-resistance mutation at RT position 65 [55].

In the six trials, 153 (10.6%) of the 1428 participants developed VF by week 24 (ARTIST), 48 (SAILING, DAWNING, and IMPAACT P1093), or 96 (NADIA and ODYSSEY). The median proportion with VF was 13.0%, ranging from 3.5% to 25.3%. Overall, 113 (7.9% of the total) had samples undergoing GRT at time of VF. Among these, 23 (1.6% of the total, or 20.4% of those tested) had samples containing one or more major non-polymorphic INSTI DRMs. Additionally, seven participants (in SAILING, DAWNING, and IMPAACT P1093) were later identified with emergent INSTI DRMs at subsequent time points, specifically weeks 60, 72 (two individuals), 108, 120, and 168 (two individuals) [52,54,62].

Among the 30 individuals that eventually developed INSTI DRMs, the patterns of DRMs were R263K alone (n=8), G118R alone (n=5), N155H alone (n=2), Q148R/K alone (n=2), R263K+G118R, R263K+S230R, G118R+H51Y+E138K, G118R+T66I, G118R+E92Q, G118R+T66I+E138K, G118R+T66A+E138K, G118R+T66I+E138K+G149A, G118R+T66A+E138K+G149A, G118R+E138K, G118R+E138A+G140A, Q148H+N155H+E138K+G140S, and Q148R+E138A+G140A. Complete integrase sequences were available only for the 12 sequences in the DAWNING and IMPAACT trials.

Of the nine participants with emergent INSTI DRMs in the NADIA trial, six had been randomized to zidovudine (AZT) plus 3TC while three had been randomized to TDF plus 3TC. The three TDF/3TC recipients with emergent INSTI DRMs each acquired R263K, which is associated with approximately two-fold reduced DTG susceptibility [3]. In contrast, the six AZT/3TC recipients included five who acquired multiple mutations including G118R or Q148R that are predicted to be associated with a high level of reduced DTG susceptibility.

Two additional clinical trials of DTG-containing ART in adults with VF on an NNRTI-containing regimen were reported at scientific meetings. In the VISEND trial, 10% of 208 PLWH treated with DTG plus TDF/3TC and 14% treated with DTG plus tenofovir alafenamide (TAF)/FTC experienced VF at week 48 [65]. In the D2EFT trial, 22% of 291 PLWH treated with DTG plus TDF/3TC or TDF/FTC experienced VF at week 48 [66]. Data on emergent INSTI DRMs in these trials have yet to be reported.

Five cohort studies described the use of DTG plus two NRTIs in PLWH with a history of VF on an NNRTI-containing regimen (Supplementary Table 3). Two were conducted in upper-income countries [67,68] and three in Sub-Saharan Africa [69–71]. The two upper-income country cohorts included 374 individuals with variable ART histories of whom approximately 70% were virologically suppressed prior to receiving DTG. Seven (1.9% of the total) individuals underwent GRT at the time of VF, and two developed the INSTI DRM R263K.

The three African cohorts included two cohorts containing 3117 individuals of whom approximately 95% were virologically suppressed on their first-line NNRTI-containing regimen [69,70] and one cohort of 139 individuals of whom just 10% were virologically suppressed [71]. In two cohorts, the proportions of those who were not suppressed at baseline and who developed VF were estimated to be 8% in one study and 10% in the other study [69,71]. GRT was performed in at least 20 individuals in the three cohorts. Two individuals with baseline resistance to TDF and 3TC developed INSTI DRMs – R263K in one individual and G118R in another [69].

Five cross-sectional studies from Sub-Saharan Africa examined the prevalence of INSTI DRMs in PLWH with VF while receiving DTG plus two NRTIs (Supplementary Table 3). The populations in these studies included mixtures of individuals with different ART histories and different proportions with VS prior to receiving DTG. The proportions of individuals with VF undergoing GRT also varied between studies. In a study from Malawi, GRT was performed on 27 samples from more than 6400 individuals with VF [72]. In a study from Nigeria, GRT was performed on 33 samples from 281 individuals with VF [73]. In three studies from Tanzania, GRT was performed in nearly all of the 181 individuals with VF [74–76]. Among the estimated 214 individuals in these five studies undergoing GRT, 19 (8.9%) developed an INSTI DRM – most commonly G118R and R263K.

### ART-Experienced PLWH with VS

#### DTG plus 2 NRTIs

Three clinical trials examined the efficacy of DTG plus two NRTIs in 877 ART-experienced PLWH with VS for six or more months [77–80] (Table 4). The NEAT022 and STRIIVING trials enrolled individuals with no history of VF or NRTI-associated DRMs. By contrast, the 2SD trial enrolled individuals who were on a second-line ritonavir-boosted protease inhibitor-containing regimen. These individuals were therefore likely to have harbored a high proportion of viruses with NRTI-resistance DRMs as a result of VF on their previous first-line regimen.

**Table 4.**
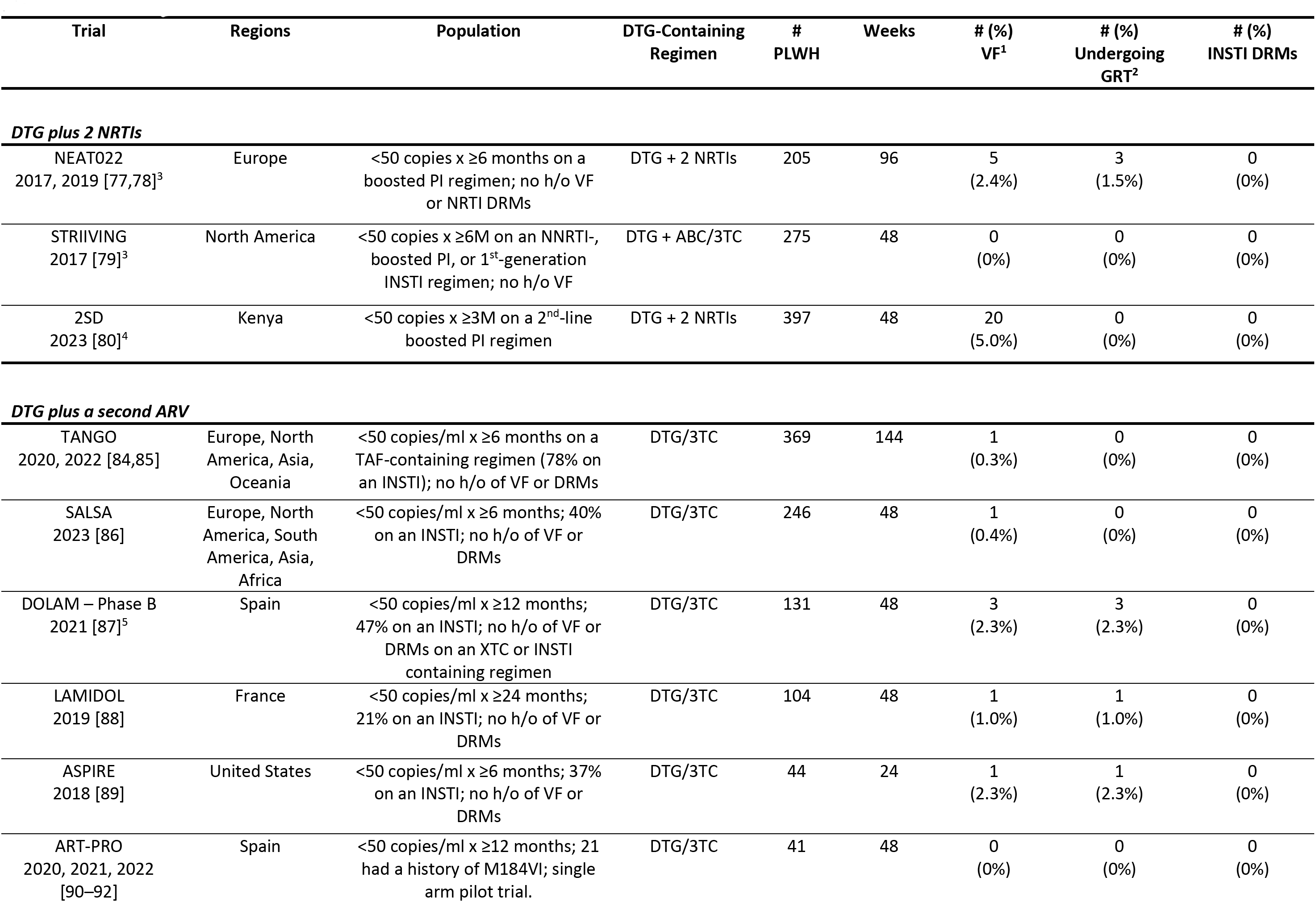
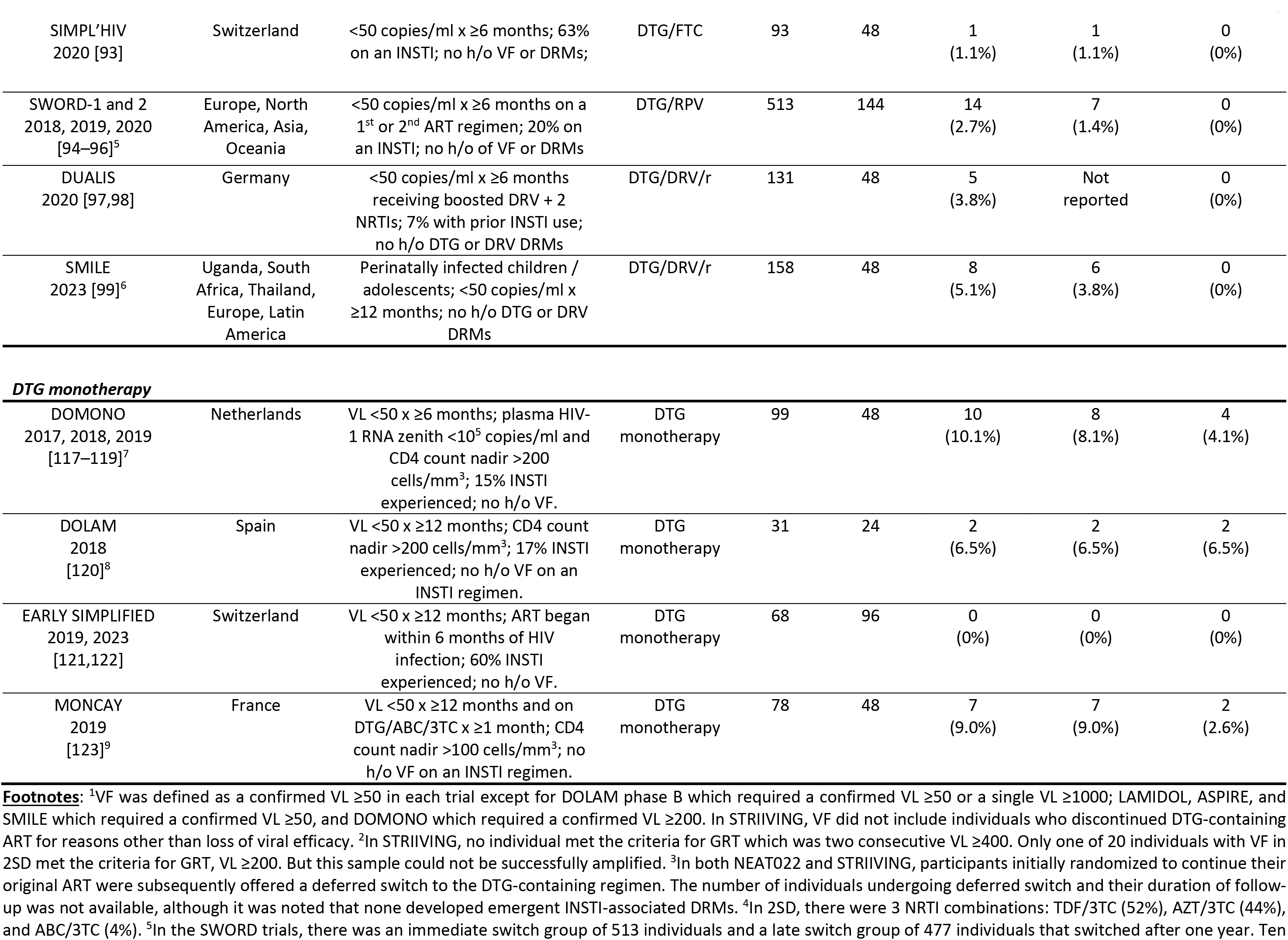
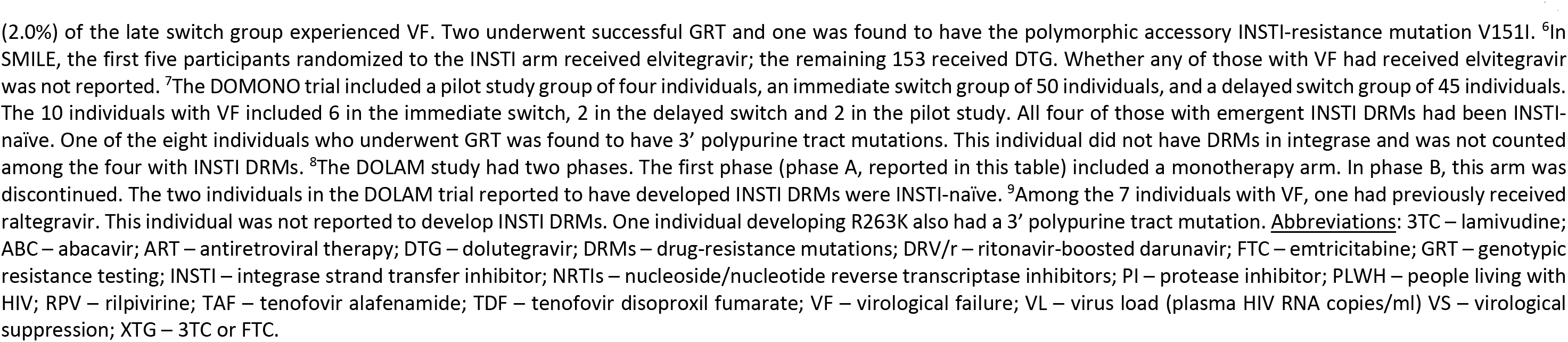
Virological Failure (VF) and Prevalence of Emergent INSTI-Associated DRMs in Clinical Trials of ART-Experienced PLWH with Virological Suppression (VS) Receiving a DTG-Containing Regimen.

The proportion of individuals with VF defined as a confirmed plasma HIV-1 RNA level ≥50 copies/ml by week 48 in STRIIVING and 2SD, and by week 96 in NEAT022 ranged from 0% to 5.0%. Three individuals in NEAT022 and none in STRIIVING or 2SD underwent GRT. In the 2SD trial, 19 of the 20 individuals with VF had a plasma HIV-1 RNA level below 200 copies/ml, and virus amplification was unsuccessful in the one individual with an RNA level above 200 copies/ml.

Three European cohort studies described 2204 PLWH with VS for six or more months on a wide variety of past ART regimens who changed their therapy to DTG plus two NRTIs [81–83] (Supplementary Table 4). The proportion with a history of VF prior to VS ranged from 11% to 80%. The proportion with M184VI on a historical GRT ranged from 8% to 80%. VF was reported in 52 (2.4%) individuals after a median of 9 to 21 months. A history of M184VI prior to starting DTG-containing ART was not reported to be associated with an increased risk of VF in any study. Of 15 individuals with VF undergoing GRT, none developed INSTI DRMs.

#### DTG Plus One Additional ARV

Ten clinical trials examined the efficacy of a two-drug DTG-containing regimen in PLWH with VS for six or more months including six trials of DTG/3TC [84–92], one trial of DTG/FTC [93], one trial of DTG/rilpivirine [94–96], and two trials of DTG plus ritonavir-boosted darunavir (DRV/r) [97–99] (Table 4). Among the seven trials of DTG/3TC or DTG/FTC, six enrolling 987 individuals excluded those with a history of VF or major DRMs. In these six trials, 21% to 78% of participants had been on an INSTI prior to starting dual therapy, 8 (0.8%) experienced VF and six (0.6%) underwent GRT, but none developed INSTI DRMs. One pilot trial of 41 individuals stratified its enrollment to include 21 individuals with a history of M184VI [90–92]. None of the participants in this trial had a history of receiving an INSTI. By week 96, the four individuals with VF in this trial included three with a history of M184VI. None of the four individuals developed emergent INSTI DRMs.

Among the 513 SWORD trial participants randomized to switch to DTG/RPV, the cumulative proportion with VF was 2.7% at week 144. Seven individuals met the criteria for GRT and none developed INSTI DRMs. At least four individuals acquired NNRTI-associated DRMs. Among the 477 control participants who underwent a deferred switch to DTG/rilpivirine, 10 (2.1%) experienced VF by week 48. The one individual undergoing GRT did not acquire INSTI DRMs.

DUALIS and SMILE assessed the efficacy of DTG plus DRV/r at maintaining VS in PLWH with diverse ART histories before attaining VS. SMILE was a study of predominantly perinatally infected children and adolescents. The prevalence of VF at week 48 was 3.8% in DUALIS and 5.1% in SMILE. Six SMILE participants underwent GRT. No participant in either trial acquired an INSTI DRM.

Eighteen observational studies described 5930 PLWH with VS for six or more months on a wide variety of previous ART regimens who changed their therapy primarily to DTG/3TC or DTG/rilpivirine (Supplementary Table 5) [83,100–116]. Sixteen studies were from Western Europe, one was from Turkey, and one was from South Korea. The median proportion with a history of VF prior to starting DTG was 25% in the eight studies reporting this information. The median proportion with a history of M184VI was 4.1% in the six studies reporting this information. VF, which was usually defined as a confirmed plasma HIV-1 RNA level ≥50 copies/ml or a single RNA level ≥1000 copies/ml, was reported in 203 (2.9%) individuals after a median of 12 months. Of the 41 PLWH undergoing GRT in eleven of the studies, three developed emergent INSTI DRMs including R263K; G118R and R263K; and T66A, G118R, and E138K [111,113,115].

#### DTG monotherapy

Table 4 also summarizes data from four clinical trials of DTG monotherapy that enrolled 276 PLWH with VS for six or more months [117–123]. The proportion with a history of receiving an INSTI ranged from 15% and 17% in DOMONO and DOLAM to 60% in EARLY SIMPLIFIED and 100% in MONCAY. Participants in DOMONO and EARLY SIMPLIFIED were required to have no history of VF, while those in DOLAM and MONCAY were required to have no history of VF on an INSTI-containing regimen. Participants in EARLY SIMPLIFIED were required to have initiated ART within six months from their primary HIV-1 infection. VF was defined as a confirmed plasma HIV-1 RNA level ≥200 copies/ml in DOMONO and ≥50 copies/ml in the remaining trials.

Overall, 19 (6.9%) of 276 individuals experienced VF by week 24 or 48, 17 (6.2%) underwent GRT, and eight (2.9%) developed one or more INSTI DRMs. INSTI DRMs emerged in one or more participants in each of the trials except for EARLY SIMPLIFIED. The eight individuals with emergent INSTI DRMs acquired the following mutations: R263K (n=2), N155H, S230R, N155H+E92Q, N155H+S147G, N155H+E138K+G140S, and N155H+Q148R+S147G. Complete integrase sequences were not available for any of the samples undergoing GRT.

One of the eight individuals who underwent GRT in the DOMONO trial was reported to have developed multiple 3’ polypurine tract mutations without DRMs in the integrase gene. One of the seven individuals who underwent GRT in the MONCAY trial was found to have a single 3’ polypurine tract mutation in combination with R263K.

Three small European observational studies described 123 PLWH with VS for six or more months who were treated with DTG monotherapy (Supplementary Table 6) [124–126]. Within 24 months, five cases of VF and emergent INSTI DRMs were reported.

## DISCUSSION

Estimating the prevalence of VF with emergent INSTI DRMs in PLWH receiving a DTG-containing regimen is crucial for updating HIV-1 treatment guidelines, developing strategies to monitor acquired DTG resistance, and determining potential indications for GRT in LMICs, where GRT is not routinely available or recommended. Three of the six clinical scenarios addressed in this review are relevant to the global expansion of DTG-containing regimens in LMICs: ART-naïve PLWH receiving a first-line regimen containing DTG plus two NRTIs (scenario 1); ART-experienced PLWH with VF on a first-line NNRTI-containing regimen switching to a second-line regimen comprising DTG plus two NRTIs (scenario 3); and ART-experienced PLWH with VS switching to a regimen containing DTG plus two NRTIs (scenario 4). In contrast, the use of two-drug regimens for initial ART (scenario 2) or maintenance ART for PLWH with VS (scenario 5) is recommended only in upper-income countries while DTG monotherapy (scenario 6) is not recommended in any scenario outside of a clinical trial.

Of the three scenarios relevant to LMICs, a discernable risk of VF and emergent INSTI DRMs was observed only in clinical scenario 3. The prevalence of VF and emergent DRMs in this scenario, at 1.6% over a period of 48 to 96 weeks in clinical trials, was lower than anticipated considering that most participants probably had viruses resistant to 3TC and that a significant proportion likely had viruses with reduced TDF susceptibility. However, in three of these trials, a continued risk of VF with emergent INSTI DRMs was observed in the second and third year of follow-up [52,54,62]. Preliminary observations based on limited data from these trials suggest that the risk of VF and emergent INSTI DRMs may be higher in children compared with adults and in those receiving AZT/3TC compared with those receiving TDF/3TC. Further studies are required to identify potential factors that influence the risk of VF and emergent INSTI DRMs in this scenario.

In two scenarios, we were able to estimate the prevalence of INSTI DRMs in the subset of individuals developing VF and undergoing GRT. Among ART-naïve PLWH receiving a first-line regimen of DTG plus two NRTIs, 1 (0.7%) of 147 with VF undergoing GRT developed INSTI DRMs. This is much lower than the prevalence of NNRTI-resistance mutations in PLWH with VF on a first-line NNRTI-containing regimen. For example, combined 48 and 96 week data from the SINGLE [11], ADVANCE [16], and NAMSAL [19] trials demonstrated that 25 (58.0%) of 43 PLWH with VF on a first-line EFV-containing regimen developed NNRTI-associated DRMs. The infrequent detection of INSTI DRMs in PLWH with VF on a first-line DTG-containing regimen suggests that incomplete adherence rather than emergent drug resistance is a more significant contributor to VF in those receiving a first-line DTG-containing regimen compared with a first-line NNRTI-containing regimen. This suggests that in PLWH with confirmed VF on a first-line DTG-containing regimen enhancing adherence support may prove more beneficial than transitioning to a second-line ART regimen. In contrast, among ART-experienced PLWH receiving a second-line regimen of DTG plus two NRTIs, 23 (20.4%) of 113 individuals with VF undergoing GRT developed INSTI DRMs suggesting a need to investigate a potential role of GRT in this setting to inform treatment changes.

The 43 clinical trials analyzed in this review described populations that were well defined in terms of their ART history, plasma HIV-1 RNA level at the time their DTG-containing regimen was initiated, and the ARVs co-administered with DTG. These trials also uniformly reported the proportion of individuals with VF, the proportion undergoing GRT, and the proportion with emergent INSTI DRMs at specific time points. However, clinical trials may report lower estimates of VF with emergent drug resistance because trial participants are more likely to be adherent to ART than PLWH in real-world settings and because in a trial, GRT is performed promptly following the detection of VF, reducing the time HIV replicates while an individual is receiving suboptimal therapy. Moreover, participants in a clinical trial of first-line ART are usually required to be ART-naïve. In contrast, in real-world settings, a significant proportion of individuals presenting for first-line ART are re-initiating therapy and may harbor acquired NRTI-associated DRMs [127–129].

Observational cohorts theoretically provide a more cost-effective approach for estimating the prevalence of VF with emergent INSTI DRMs in real-world settings. However, we found that, in practice, few published cohort studies contributed relevant quantitative data because all but four were from upper-income countries where ART histories were highly variable and where decisions about whether to switch to a DTG-containing regimen and ARVs to administer with DTG were highly individualized.

Cross-sectional studies in which GRT is performed in persons with confirmed VF on a DTG-containing regimen provide an alternate mechanism for assessing the risk of emergent DTG resistance. We described six such studies including one from Brazil of 113 PLWH with confirmed VF on a first-line regimen of DTG plus two NRTIs [42] and five from Sub-Saharan Africa. The studies from Sub-Saharan Africa included approximately 214 individuals most of whom had prior ART experience before initiating DTG treatment [72–76]. The study from Brazil found that approximately six percent of the samples tested were found to have INSTI DRMs while in the studies from Sub-Saharan Africa, approximately nine percent of the samples tested had INSTI DRMs. However, in these studies, either the total number of persons receiving DTG-containing ART or their past ART histories were unknown making it impossible to estimate the prevalence of VF with emergent INSTI DRMs in any particular clinical scenario.

With the rising global adoption of DTG-based antiretroviral therapy (ART) and prolonged treatment durations for PLWH, the development of uniform protocols to surveil for acquired DTG resistance is crucial. The WHO has devised several survey models [130], which have been complemented by additional methodologies implemented by the U.S. Centers for Disease Control and Prevention (CDC) [131–135]. These cost-effective, cross-sectional studies focus on PLWH who with VF during DTG-based ART, as opposed to more resource-intensive longitudinal cohort studies. Recent presentations at a scientific meeting highlighted four CDC collaborative studies in Sub-Saharan Africa [132–135]. The number of samples tested per study ranged from 65 to 512 and the proportion of samples with an INSTI DRM ranged from 3% to 21%. To improve the impact of such studies, integrating estimates of the total population under treatment and comprehensive ART histories would provide a more accurate projection of resistance risk across the treated demographic.

## Funding statement

This work was funded by a grant from the National Institutes of Health: 2R24AI13661806. The funder played no role in this review.

## Competing interests statement

Robert W. Shafer has received honoraria for participation in advisory boards from Gilead Sciences and GlaxoSmithKline and speaking honoraria from Gilead Sciences and ViiV Healthcare.

## Ethics approval statement

This study does not involve human participants.

## Data Availability

All data produced in the present work are contained in the manuscript.

## Notes

### Author Declarations

The study used ONLY openly available human data that were originally located at PubMed.

