## Supplementary Materials for "Risk of Emergent Dolutegravir Resistance Mutations In People Living With HIV: A Rapid Scoping Review"

| **Table S1. Virological Failure (VF) and Prevalence of Emergent INSTI-Associated DRMs in Cohorts of ART-Naïve PLWH Receiving DTG Plus Two NRTIs** | | | | | | | |
| --- | --- | --- | --- | --- | --- | --- | --- |
| **Author / Year** | **Region/**  **Country** | **Population** | **DTG-containing regimen** | **#**  **PLWH** | **# (%)**  **with VF^1^** | **# (%)**  **undergoing GRT** | **# (%)**  **with INSTI DRMs** |
| ***Cohort studies*** | | | | | | | |
| Rhee 2020 [1] | U.S.  2014-2017 | Adults | DTG + 2 NRTIs | 355 | After a median of 26M, 6.2% had VF defined as a single VL ≥200. | 5 (1.4%) | 0 (0%) |
| Armenia 2020 [2] | Italy  2014-2017 | Adults | DTG + 2 NRTIs | 269 | At 12M, ~3.3% had VF defined as a confirmed VL ≥50. | 4 (1.5%) | 0 (0%) |
| Ntamatungiro 2023 [3] | Tanzania  2019-2020  (KIULARCO) | Adults | DTG +  TDF/3TC | 436 | At 12M, 6.9% had VF defined as a confirmed VL ≥1000. | 2 (0.5%) | 0 (0%) |
| Mondi 2019 [4] | Italy  2015-2017  (ICONA) | Adults | DTG + 2 NRTIs | 882 | After a median of 9M, ~2.6% had VF defined as a confirmed VL ≥50. | 4 (0.5%) | 0 (0%) |
| Suárez-García 2023 [5] | Spain  2018-2021  (CORIS) | Adults | DTG + 2NRTIs | 537 | At 12M, 10/339 (2.9%) had VF defined as a confirmed VL ≥50. | Not reported | 0 (0%) |
| Grayhack 2018 [6]^2^ | U.S.  2015-2018 | Pregnant women | Not reported | 66 | At delivery, 13/57 (22.8%) had VF defined as a single VL ≥20. | Not reported | 0 (0%) |
| Chin 2020 [7] | Korea  2014-2017 | Adults | DTG +  ABC/3TC | 153 | After a median of 2M, 0% had VF defined as a confirmed VL ≥200. | Not applicable | Not  applicable |
| ***Cross-sectional GRT studies of PLWH with VF on a DTG-containing regimen*** | | | | | | | |
| Diaz 2023 [8] | Brazil  2017-2018 | Adults | DTG +  TDF/3TC | Not known | Not reported | 113 | 7 (6.2%): R263K (n=4); G118R; E138A; G140R |
| **Footnotes**: ^1^The % with VF is preceded by ‘~’ when the total number of individuals evaluated included some PLWH who did not receive DTG plus 2 NRTIs, most commonly DTG plus 3TC. ^2^42.4% on DTG prior to pregnancy, 24.2% initiated ART with DTG during pregnancy, 33.3% switched to DTG during pregnancy. Abbreviations: 3TC – lamivudine; ABC – abacavir; ART – antiretroviral therapy; DRMs – drug-resistance mutations; DTG – dolutegravir; GRT – genotypic resistance testing; INSTI – integrase strand transfer inhibitor; M – months; NRTIs – nucleoside RT inhibitors; PLWH – people living with HIV; TDF – tenofovir disoproxil fumarate; VF – virological failure; VL – virus load as plasma HIV RNA copies/ml. | | | | | | | |

| **Table S2. Virological Failure (VF) and Emergent INSTI-Associated DRMs in Cohorts of ART-Naïve PLWH Receiving DTG Plus 3TC** | | | | | | |
| --- | --- | --- | --- | --- | --- | --- |
| **Author / Year^1^** | **Region / Country** | **Population^2^** | **# PLWH** | **# (%)**  **with VF**  **at 12 months^3^** | **# (%)**  **Undergoing GRT** | **# (%)**  **with**  **INSTI DRMs** |
| Suárez-García 2023 [5] | Spain  2018-2021 | Adults | 401 | 3  (0.7%) | 1  (0.2%) | 0  (0%) |
| Cabello-Ubeda 2022 [9] | Spain  ≤2020 | Adults | 135 | 1  (0.7%) | 1  (0.7%) | 0  (0%) |
| Li 2022 [10] | China  2020-2022 | Adults | 45 | 0  (0%) | Not applicable | Not applicable |
| **Footnotes:** ^1^The study by Suárez-García et. al. also described a cohort of 282 individuals receiving DTG plus TDF/FTC and 255 individuals receiving DTG plus ABC/3TC (see Table S1). ^2^Baseline GRT was performed in 165 (41.1%) of individuals in the study by Suárez-García et. al., 38 (28.1%) of individuals in the study by Cabello-Ubeda et. al., and all of the individuals in the study by Li et. al. One individual in the study by Suárez-García et. al. and two individuals in the study by Cabello-Ubeda et. al. had a baseline M184 mutation and were changed to a different regimen. No baseline NRTI DRMs were reported in the study by Li et. al. ^3^VF was defined as a confirmed VL ≥50 or a single VL ≥1000 in each study after 48 weeks. In the study by Suárez-García et. al., 22 individuals (5.5%) discontinued therapy for reasons other than author-defined VF. In the study by Cabello-Ubeda et. al., 19 individuals (14.1%) discontinued therapy for reasons other than author-defined VF or were lost to follow-up. Abbreviations: 3TC – lamivudine; ABC – abacavir; ART – antiretroviral therapy; DRMs – drug-resistance mutations; FTC – emtricitabine; GRT – genotypic resistance testing; INSTI – integrase strand transfer inhibitor; PLWH – people living with HIV; VF – virological failure; VL – virus load as plasma HIV RNA copies/ml. | | | | | | |

| **Table S3 Virological Failure (VF) and Emergent INSTI-Associated DRMs in Cohort Studies Describing the Use of DTG Plus Two NRTIs in PLWH With Previous VF Who Were Not Uniformly Virologically Suppressed at DTG Initiation** | | | | | | | | | |
| --- | --- | --- | --- | --- | --- | --- | --- | --- | --- |
| **Author / Year** | **Region / Country** | **Population** | **ART**  **History** | **Pre-Switch**  **VL** | **#**  **PLWH** | **DTG**  **ART** | **# (%)**  **with VF** | **# (%)**  **Undergoing GRT** | **# (%)**  **with INSTI**  **DRMs** |
| ***Upper-income country cohort studies*** | | | | | | | | | |
| Lepik 2017 [11]^1^ | Canada  (2014-2015) | Adults | NNRTI and/or PI-containing ART;  15% baseline NRTI DRMs. | ~70% had VL <50 | 252 | DTG +2 NRTIs | At 12M, ~17% had VF defined as a confirmed VL ≥50 or DTG discontinuation. | 3 (1.2%) | 2 (0.8%):  R263K (2) |
| Sorstedt 2018 [12]^2^ | Sweden  (≤2017) | Adults | Variable; 21% INSTI-experienced; Baseline NRTI DRMs was an inclusion criterion; 37% had M184VI. | 75% had VL <50 | 122 | DTG + 2 NRTIs | After a median of 18M, 3.3% had VF defined as a single VL ≥200. | 4 (3.3%) | 0 (0%) |
| ***Low- and middle-income country cohort studies*** | | | | | | | | | |
| Semengue 2023 [13] | Cameroon  2021 | Adults | TDF/3TC/EFV | 90% had a “detectable” VL | 139 | DTG + TDF/3TC | At 14M, 7.9% had VF defined as VL ≥1000 | 11 (7.9%) | 0 (0%) |
| Brown 2022 [14] | Lesotho  2020  (DO-REAL study) | Adults | Dual NRTI / NNRTI | Of 96% with available VL: 96% had VL <100 | 1225 | DTG + TDF/3TC | After a median of 4M, 95% had an available VL and 1% had VF defined as VL ≥1000 | 7 (0.6%) | 0 (0%) |
| Schramm 2022 [15] | Malawi  2019 (MSF program) | Adults | Dual NRTI / NNRTI | 95% had VL <50 | 1892 | DTG + TDF/3TC | At 18M, 10% (7/69) of those who were viremic before TDF/3TC/DTG developed VF defined as VL ≥50 | Not reported | 2 individuals with baseline dual TDF/3TC resistance developed INSTI DRMs: R263K; G118R |
| ***Cross-sectional GRT studies of PLWH with VF on a DTG-containing regimen*** | | | | | | | | | |
| Van Oosterhout  2022 [16]^3^ | Malawi  2020-2021 (National HIV Treatment Program) | Adults / Children / Adolescents | Naïve, 1^st^-line ART, 2^nd^-line ART | Not reported | NR | Presumed DTG + 2 NRTIs | 6462 (~8%) individuals had VF defined as a single VL ≥1000 | 27 (0.4%) of those with VF | 8 samples had INSTI DRMs: R263K (3); R263K, E157Q (2); R263K, M50I; R263K+5 accessory DRMs; H51Y, S147G |
| Abdullahi 2023 [17]^4^ | Nigeria  2021 | Adults | Naïve, 1^st^-line ART, 2^nd^-line ART | Not reported | 4263 | DTG + TDF/3TC | 281 (6.7%) had VF defined as a single VL ≥1000. | 33  (0.8%) | 1 sample had INSTI DRMs: T66A, G118R, E138K, R263K |
| Kamori 2023 [18]^5^ | Tanzania  2020 | Adults / Children | 1^st^-line ART, 2^nd^-line ART | Not reported | NR | Presumed DTG + 2 NRTIs | 84 (5.0%) had VF defined as a single VL ≥1000. | 84  (5.0%) | 4 samples had INSTI DRMs: R263K; G118R; T66A, G118R, E138K; E138K, G140A, Q148K |
| Khamadi 2023 [19]^6^ | Tanzania  2019-2021 | Children / Adolescents | 1^st^-line ART; 2^nd^-line ART | 87%  <1000 | 502 | DTG + 2 NRTIs | 64 (12.7%) of individuals had VF defined as a single VL ≥1000 | ~40  (~8.0%) | 3 samples had INSTI DRMs: R263K (3) |
| Bwire 2023 [20] | Tanzania  2023 | Adults | Naïve, 1^st^-line ART, 2^nd^-line ART | Not reported | 600 | DTG + TDF/3TC | 33 (5.5%) of individuals had VF defined as a single VL ≥1000 | 30  (5.0%) | 3 samples had INSTI DRMs: G118R, E138K; E138K, G140A, Q148K; T66I, G118R, E138K |
| Footnotes: ^1^This study also included individuals (21% of the total) who had been INSTI-experienced. The “~” indicates that the precise proportions with baseline VS and with VF following DTG were not available for the subset who were INSTI experienced. ^2^One individual had VF while receiving raltegravir prior to receiving DTG but had not developed INSTI DRMs; ^3^87 applications were submitted for sequencing; 33 samples were selected, and 27 were successfully sequenced. ^4^Sample collection was successful in 61 individuals and 33 of these were successfully sequenced. ^5^The study included 367 individuals who did not receive a DTG-containing regimen. However, because GRT was successful in 98.5% of individuals with VF in the study, we estimated that all had undergone GRT. ^6^A random selection of 707 children and adolescents on first- or second-line ART for ≥6 months were selected of whom 71.0% were receiving DTG plus 2 NRTIs. The number undergoing GRT was estimated from the 63% rate of successful sequencing for the complete cohort. Abbreviations: 1^st^-line ART – generally refers to a nonnucleoside RT inhibitor plus 2 NRTIs; 2^nd^-line ART – generally refers to a protease-inhibitor containing regimen; 3TC – lamivudine; ART – antiretroviral therapy; DRMs – drug-resistance mutations; GRT – genotypic resistance testing; INSTI – integrase strand transfer inhibitor; NR – not reported; NRTIs – nucleoside reverse transcriptase inhibitors; PLWH – people living with HIV; TDF – tenofovir disoproxil fumarate; VF – virological failure; VL – virus load as plasma HIV RNA copies/ml; VS – virological suppression. | | | | | | | | | |

| **Table S4. Virological Failure (VF) and Emergent INSTI-Associated DRMs in Cohorts of ART-Experienced PLWH with VS Receiving DTG Plus Two NRTIs** | | | | | | | |
| --- | --- | --- | --- | --- | --- | --- | --- |
| **Author / Year^1^** | **Region/**  **Country** | **ART History^2^** | **#**  **PLWH** | **DTG ART** | **# (%)**  **with VF^3^** | **# (%) Undergoing GRT** | **# (%) With INSTI DRMs** |
| Olearo 2019 [21] | Europe | VS (median >84M); 19% had h/o INSTI; >30% h/o VF;  8.4% had h/o M184VI | 1626 | DTG + ABC/3TC | 21 (1.3%)  (median 9M) | 6  (0.4%) | 0  (0%) |
| Borghetti 2022 [22] | Italy | VS (median ~48M); INSTI history not reported; 11% h/o VF;  12% h/o M184VI | 424 | DTG + ABC/3TC (72%) or TDF/FTC (28%) | 31 (7.3%)  (median 21M) | 6  (1.4%) | 0  (0%) |
| Jary 2020 [23] | France | VS (median 36M); INSTI-naïve;  ~82% had h/o VF and M184VI | 154 | DTG + ABC/3TC | 0 (0%)  (at 12M) | 3  (2.0%) | 0  (0%) |
| **Footnotes:** ^1^The study by Borghetti et. al. also included cohorts with virological suppression (VS) that received DTG/3TC and was therefore also in Table S5. ^2^M (months). With the exception of Olearo et al., each study explicitly reported that no individual experienced VF while receiving an INSTI. ^3^VF was defined as a confirmed VL ≥50 or a single VL ≥200 for Jary et. al., and Borghetti et. al. and as a confirmed VL ≥50 or as single VL ≥50 at the last time point for Olearo et. al. M184VI did not appear to increase the risk of VF in the study by Olearo et. al. Abbreviations: 3TC – lamivudine; ABC – abacavir; ART – antiretroviral therapy; DRMs – drug-resistance mutations; DTG – dolutegravir; FTC – emtricitabine; GRT – genotypic resistance testing; h/o – history of; INSTI – integrase strand transfer inhibitor; M – months; PLWH – people living with HIV; TDF – tenofovir disoproxil fumarate; VF – virological failure; VS – virological suppression. | | | | | | | |

| **Table S5. Virological Failure (VF) and Emergent INSTI-Associated DRMs in Cohorts of ART-Experienced PLWH with VS Receiving DTG Plus a 2^nd^ ARV** | | | | | | | |
| --- | --- | --- | --- | --- | --- | --- | --- |
| **Author/Year^1^** | **Countries / Years** | **ART**  **History^2^** | **#**  **PLWH** | **DTG**  **ART** | **# (%)**  **with VF^3^** | **# (%) Undergoing GRT** | **# (%)**  **with INSTI**  **DRMs** |
| Palmier 2022 [24] | Spain  2020-2019 | VS (median duration not reported); 27.1% had h/o VF; 70.7% INSTI-experienced; 4.7% had historical M184VI. | 358 | DTG/3TC | After a mean of 36M, 3.6% had VF defined as VL ≥50. | 9  (2.5%) | 1 (0.3%);  R263K |
| Bowman 2023 [25]^4^ | UK  2015-2021 | VS (median duration not reported); 3.7% had VL ≥50, % with h/o VF not reported; 14.3% INSTI-experienced. | 552 | DTG/XTC (86.6%)  DTG/RPV (13.4%) | After a median of 11M, 5/460 (1.1%) on DTG/3TC had VF. After a median of 28M, 1/74 (1.4%) on DTG/RPV had VF. VF was defined as a confirmed VL ≥200. | 5  (0.9%) | 1 (0.2%) individual on DTG/3TC: T66A, G118R, E138K |
| Knobel 2023 [26]^5^ | Spain  2019-2022 | VS (median duration not reported); 42% had been on a DTG-containing regimen; 12% had received a first-generation INSTI; % with h/o VF not reported. | 358 | DTG/3TC | After a median of ~36M, 1.1% had VF defined as a confirmed VL ≥200. | 4  (1.1%) | 1 (0.3%);  G118R, R263K |
| Baldin 2019 [27] | Italy  (≤2019) | VS (median 96M); 43.4% had h/o VF; 19.5% INSTI-experienced; 9.0% had historical M184VI. | 221 | DTG/3TC | After a median of 25M, 3.2% had VF defined as a confirmed VL ≥50 or a single VL ≥1000. | 7  (3.2%) | 0  (0%) |
| Borghetti 2022 [22] | Italy  2014-2020 | VS (median 48M); 10.8% had h/o VF; INSTI history not reported;  3.9% had historical M184VI. | 204 | DTG/3TC | After a median of 20M, 4.9% had VF defined as a confirmed VL ≥50 or a single VL ≥200. | 1  (0.5%) | (0%) |
| Calza 2020 [28] | Italy  2016-2018 | VS (median 41M); No h/o VF on INSTI or XTC-containing regimen;  80.0% INSTI-experienced. | 59 | DTG/3TC | At 12M, 3.4% had VF defined as a single VL ≥20. | 2  (3.4%) | 0  (0%) |
| Calza 2023 [29] | Italy  2018-2020 | Age > 65; VS (median 71M); No h/o VF; INSTI-naïve. | 72 | DTG/3TC | At 12M, 4.2% had VF defined as a single VL ≥20. | 3  (4.2%) | 0  (0%) |
| Ciccullo 2021 [30] | Italy  2016-2021 | VS (median 29M); 22.3% had h/o VF; 27.5% INSTI-experienced; 4.2% had historical M184VI. | 785 | DTG/3TC | After a mean of 30M, 2.3% had VF defined as a confirmed VL ≥50 or a single VL ≥1000. | Not reported | 0  (0%) |
| Ciccullo 2023 [31] | Italy  2015-2021 | VS (median 80M); 32.7% had h/o VF; 27.3% INSTI-experienced. | 592 | DTG/3TC (51.7%)  DTG/RPV (48.3%) | After a median of 25M, 2.6% on DTG/3TC and after a median of 28M, 1.0% on DTG/RPV had VF defined as a confirmed VL ≥50 or a single VL ≥1000. | 11  (0.2%) | 0  (0%) |
| Maggiolo 2022 [32] | Italy  2015-2017 | VS (median 75M); % with h/o VF not reported; 22.5% INSTI-experienced;  0% had historical M184VI. | 218 | DTG/3TC | After a mean of 64M, 0% had VF defined as a confirmed VL ≥50. | Not applicable | Not  applicable |
| Lee 2022 [33] | Korea  2020-2022 | VS (median duration not reported); % with h/o VF not reported; 91% INSTI-experienced; 2.0% had historical M184VI. | 131 | DTG/3TC | At 12M, 0% had VF defined as a VL ≥1000. | Not applicable | Not  applicable |
| Ergen 2022 [34] | Turkey  2016-2021 | VS (median duration not reported); No h/o VF; 80.9% INSTI-experienced. | 63 | DTG/3TC | After a median of 10M, 0% had VF defined as VL ≥200. | Not applicable | Not  applicable |
| Buzon 2023 [35] | Spain  2020-2021 | VS (median duration not reported); % with h/o VF not reported; 44.5% INSTI-experienced. | 1032 | DTG/3TC | At 12M, 2.5% of 763 had VF defined as a single VL ≥50. | Not reported | 0  (0%) |
| Troya 2022 [36] | Spain  2018-2019  (DORIPEX) | VS (median duration not reported); % with h/o VF not reported; 41.0% INSTI-experienced. | 524 | DTG/RPV | At 12M, 0.6% had VF defined as a single VL ≥50. | Not reported | 0  (0%) |
| Gantner 2017 [37] | France  2014-2015 | VS (median 120M); 52% had h/o VF; 59% INSTI-experienced. | 152 | DTG/RPV | After a median of 9M, 2.0% had VF defined as a confirmed VL ≥50 or a single VL ≥1000. | 2  (1.3%) | 0  (0%) |
| Casado 2019 [38] | Spain  2015-2017 | VS (median 52M); % with h/o VF not reported; 15% INSTI-experienced. | 102 | DTG/RPV | At 12M, 1.0% had VF defined as a confirmed VL ≥50. | 1  (1.0%) | 0  (0%) |
| Poliseno 2023 [39] | Italy  2020-2021 | VS (median 96M); 66% INSTI-experienced; 6 individuals (13%) had detectable VL at the time DTG/DOR was begun | 43 | DTG/DOR | At 12M, 2.3% had VF defined as a confirmed VL ≥50 or a single VL ≥200. | 1  (2.3%) | 0  (0%) |
| Castagna 2019 [40] | Italy  2014-2018 | VS (median 44M); % with h/o VF not reported; 49.1% INSTI-experienced | 151 | DTG/ATV | After a median of 15M, 1.3% had VF defined as a confirmed VL ≥50. | 1  (0.7%) | 0  (0%) |
| **Footnotes:** ^1^Borghetti et. al. also included a cohort of individuals with VS that received DTG plus 2 NRTIs and is summarized in Table S4. Part of the Maggiolo cohort was reported in an earlier publication [41]. ^2^INSTI-experienced includes persons who received either a first-generation or second-generation INSTI. ^3^Higher rate of VF (but not INSTI DRMs) occurred in individuals with baseline M184VI in certain cohorts. In Ciccullo et. al., 2021, this included the subgroup with VS for less than 88 months. In Baldin et. al., this included the subgroup with VS for less than 96 months. In Santoro et. al., this included those with VS for less than 42 months. ^4^Included two individuals with baseline INSTI DRMs (F121Y, N155H), neither of whom developed VF; ^5^The one individual who developed INSTI DRMs had a history of receiving an elvitegravir-containing regimen but had not developed VF on that regimen. Abbreviations: 3TC – lamivudine; ABC – abacavir; ART – antiretroviral therapy; ARV – antiretroviral; ATV – atazanavir; DOR – doravirine; DRMs – drug-resistance mutations; DTG – dolutegravir; FTC – emtricitabine; GRT – genotypic resistance testing; h/o – history of; INSTI – integrase strand transfer inhibitor; M – months; PLWH – people living with HIV; RPV – rilpivirine; TDF – tenofovir disoproxil fumarate; VF – virological failure; VS – virological suppression; XTC – 3TC or FTC. | | | | | | | |

| **Table S6.**­­ **Virological Failure (VF) and Emergent INSTI-Associated DRMs in Cohorts of ART-Experienced PLWH with VS Receiving DTG Monotherapy** | | | | | | |
| --- | --- | --- | --- | --- | --- | --- |
| **Author/Year^1^** | **Countries / Years** | **ART**  **History** | **#**  **PLWH** | **# (%)**  **VF** | **# (%) Undergoing GRT at VF** | **# (%)**  **with INSTI**  **DRMs^2^** |
| Rojas 2016 [42] | Spain  2014-2015 | VS (median 96M); received a median 9 different ART regimens; 6% INSTI-experienced; no history of VF on an INSTI regimen. | 31 | At 6M, 1 individual (3.2%) had VF defined as a confirmed VL ≥50. | 1  (3.2%) | 1  (32%);  G118R |
| Oldenbuettal 2017 [43] | Germany  2014-2016 | VS (median duration not reported); 61% INSTI-experienced; no history of VF on an INSTI regimen. | 31 | At 6M, 1 individual (3.2%) had VF defined as a confirmed VL ≥50. | 1  (3.2%) | 1  (3.2%);  G140S, Q148H |
| Tebano 2020 [44] | France  2014-2018 | VS (median duration 70M); 39% INSTI-experienced; no history of VF on an INSTI regimen. | 61 | After a median duration of 24M, 4.9% had VF defined as a confirmed VL ≥50 or a single VL ≥200. | 3  (4.9%) | 3 (4.9%)  (4.9%); E138K, G140A, Q148R; E92Q; N155H |
| **Footnotes:** ^1^Tebano et. al. describes the longer term follow-up of a cohort originally reported by Katalama et. al. in 2016 [45]. Two additional cohorts included less than 30 individuals of whom none developed INSTI DRMs [46,47]. ^2^In the study by Rojas et. al., G118R was detected by next-generation sequencing of a peripheral blood mononuclear sequence in 7% of sequence reads. Abbreviations: ART – antiretroviral therapy; DRMs – drug-resistance mutations; DTG – dolutegravir; GRT – genotypic resistance testing; INSTI – integrase strand transfer inhibitor; M – months; VF – virological failure; VL – virus load as plasma HIV RNA copies/ml; VS – virological suppression. | | | | | | |
